# Transient Reductions in Postoperative Pain and Anxiety using Virtual Reality in Children

**DOI:** 10.1101/2020.09.18.20192724

**Authors:** Vanessa A. Olbrecht, Keith T. O’Conor, Sara E. Williams, Chloe O. Boehmer, Gilbert W. Marchant, Susan M. Glynn, Kristie J. Geisler, Hannah M. Pickerill, Lili Ding, Gang Yang, Christopher D. King

**Author notes:** **Address correspondence to:** Vanessa A. Olbrecht, MD, MBA Cincinnati Children’s Hospital Medical Center, 3333 Burnet Avenue, MLC 2001 Cincinnati, OH, 45229.

## Abstract

**Objective:** Virtual reality (VR) is a promising method to manage pain. Distraction-based VR (VR-D) is thought to reduce pain by redirecting attention. While VR-D can reduce pain associated with acutely painful procedures, it is unclear if VR-D can reduce pain after surgery. We assessed the ability of a single VR-D session to decrease acute postoperative pain and anxiety and explored if pain catastrophizing and anxiety sensitivity influenced the ability of VR-D to reduce these outcomes in children following surgery.

**Design:** Single-center, prospective, pilot study

**Setting:** Cincinnati Children’s Hospital Medical Center (CCHMC)

**Subjects:** 50 children/adolescents (age 7-21 years) with postoperative pain followed by the Acute Pain Service

**Methods:** Patients received a single VR-D session following surgery. Prior to the VR-D session, patients completed pain catastrophizing (PCS-C) and anxiety sensitivity (CASI) questionnaires. Primary outcome consisted of changes in pain intensity following VR-D (immediately, 15, and 30 minutes). Secondary outcomes included changes in pain unpleasantness and anxiety.

**Results:** VR-D decreased pain intensity immediately and 15-minutes after VR-D. Reductions in pain unpleasantness were observed up to 30 minutes following VR-D. Anxiety was also reduced immediately and at 15-minutes following VR-D. While patients with higher pain catastrophizing had higher baseline pain intensity and unpleasantness, they did not show larger pain reductions following VR-D compared to those with lower pain catastrophizing.

**Conclusions:** VR-D is beneficial in transiently reducing pain intensity, unpleasantness, and anxiety in children with acute postoperative pain. This study informs design of larger, controlled study assessing VR-D for acute postoperative pain and anxiety.

## 1. Introduction

Despite improved pain management strategies, the number of patients experiencing severe pain after surgery has not changed significantly over the last 20 years^1, 2^. Ineffective management of postoperative pain has consistently been shown to lead to poorer surgical outcomes^3, 4^, including delayed wound healing, development of negative pain perception, and an increased likelihood of the development of chronic pain^5, 6^. In addition, children and adolescents with acute and chronic pain are at particular risk of opioid abuse^7^, and many are exposed to narcotics initially prescribed to treat pain^8^. Surgery is a time of risk for children as the point of initial opioid exposure^9-11^. Just five days of opioid use can increase the risk of persistent use, and use for more than 8 days may increase the risk to as much as 13.5%^12^. Therefore, it is important to consider alternative, innovative strategies to provide safe, effective, nonpharmacologic therapies to children and adolescents after surgery to optimize pain management while minimizing opioid use^13, 14^.

Virtual reality (VR) has the potential to help optimize pain management. VR technology provides an immersive, multisensory, three-dimensional (3D) environment that enables individuals to have modified experiences of reality by creating a sense of “presence” for that individual, making it an excellent candidate for distraction-based therapy^15^. VR has been used to help treat anxiety disorders; control pain; support physical rehabilitation; and to distract patients during wound care, acutely painful procedures, and labor^15-27^. Furthermore, distraction-based VR (VR-D) has been shown to provide effective analgesia when combined with opioids^28^. A recent study assessed the use of VR compared to distraction-based “health and wellness” television programming - VR was more effective at reducing pain compared to the non-VR condition^29^. This was particularly true for patients with severe pain (pain scores ≥ 7 on the Numerical Rating Scale, NRS), with significant reductions in pain occurring immediately and after 2 to 3 days of use^29^. This intervention is thought to reduce pain by redirecting attention (e.g., distraction^15-27, 30-42^). While many studies have shown transient reductions in pain using VR, no study to date has focused on utilizing VR-D as part of multimodal analgesia to manage acute postoperative pain and anxiety in a pediatric population.

We performed a prospective pilot study to determine the ability of VR-D to decrease acute postoperative pain intensity (primary outcome) and whether trait pain catastrophizing^43-45^ or anxiety sensitivity^37^ influence the efficacy of VR-D to reduce pain. We also explored the impact of VR-D on levels of pain unpleasantness and anxiety (secondary outcomes) and associations with trait measures. Pain intensity is commonly assessed clinically with numerical rating scales (NRS^46, 47^) while pain unpleasantness and anxiety NRS, which may provide additional domains associated with surgery, are not extensively used clinically. We hypothesized that a single VR-D session would reduce pain intensity, pain unpleasantness, and anxiety; and that reductions in pain intensity would be greatest in patients with high levels of baseline trait pain catastrophizing and anxiety sensitivity compared to patients with lower levels of pain catastrophizing.

## 2. Methods

This single-center, prospective, pilot study assessed the ability of VR-D to transiently decrease acute postoperative pain and anxiety after a single session and determined whether pain catastrophizing and anxiety sensitivity influence the efficacy of VR-D to help pain in a broad, pediatric population with significant pain after surgery. Figure 1 summarizes the study design.

**Figure 1.**
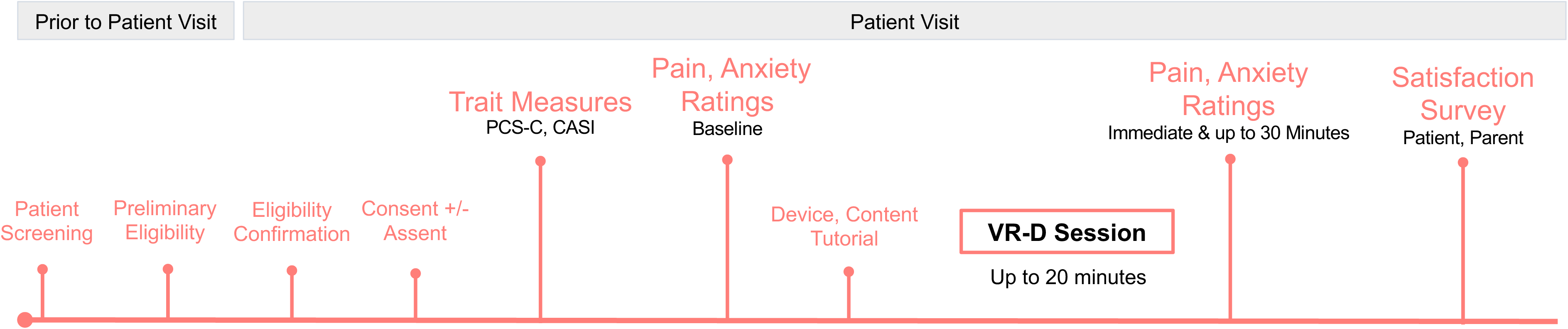
Flow Diagram of Current Study.

### 2.1 Patients

This trial was conducted in 50 children and adolescents who underwent surgery and were followed postoperatively by the Acute Pain Service at Cincinnati Children’s Hospital Medical Center (CCHMC). CCHMC is a tertiary care, academic, pediatric hospital. Patients were prospectively recruited over a 7-month period (November 2018 to July 2019). This study was approved by the Cincinnati Children’s Hospital Medical Center Institutional Review Board (IRB #2018-2892). This study was conducted in accordance with the rules and regulations applicable to the conduct of ethical research. Written consent (and assent for patients ≥ 11 years) was obtained for patients under the age of 18 from the appropriate parent or legal guardian and consent was obtained for adolescents 18 years of age and older.

#### 2.1.1 Inclusion Criteria

Patients were eligible for the study if they were between the ages of 7 and 21 years; if they were able to read, speak, and write English; and if they had major surgery resulting in significant postoperative pain necessitating care by the Acute Pain Service.

#### 2.1.2 Exclusion Criteria

Patients were excluded if they did not fall within the age criteria; if they did not speak, understand, or read English; if they had a history of developmental delay, neurological conditions (specifically epilepsy or seizure disorder, vertigo, dizziness, or significant motion sickness/nausea/vomiting), or uncontrolled psychiatric conditions; or if they had surgery of the head or neck that precluded them from wearing the VR headset.

#### 2.1.3 Patient information

Information about the patient’s age, sex, race, surgery type, and their ASA (American Society of Anesthesiologists) status, https://www.asahq.org/standards-and-guidelines/asa-physical-status-classification-system) was collected prior to the VR experience.

### 2.2 Measures

The primary outcome measure was change in pain intensity after VR-D. Secondary outcome measures included changes in pain unpleasantness and anxiety. These measures are described below.

#### 2.2.1 Pain Intensity, Pain Unpleasantness, and Anxiety

Pain intensity (Cronbach’s α = 0.93), pain unpleasantness (Cronbach’s α = 0.92), and anxiety (Cronbach’s α = 0.91) were measured using the Numerical Rating Scale (NRS)^46, 47^ before and following VR-D. A standardized approach was used to explain the difference between pain intensity and pain unpleasantness. In the scripted information given to patients, pain was described as analogous to listening to the radio, where pain intensity was related to the volume of the music playing and pain unpleasantness was related to how much the individual disliked what was playing^48^. For each of these measures, patients were asked to give a number on a scale of 0 to 10 to rate the severity of their symptom, with 0 being non-existent and 10 being the most severe pain or anxiety possible.

#### 2.2.2 Pain Catastrophizing

The Pain Catastrophizing Scale for Children (PCS-C) is a validated, self-reported measure of a child’s tendency to catastrophize about pain^49, 50^. Children rate 13 items assessing rumination, magnification, and helplessness related to thoughts about pain on a 5-point Likert scale. PCS-C summary scores are interpreted as low (0 to 14), moderate (15 to 25), and high (≥26)^51^. The PCS-C was used to determine if trait levels of catastrophizing impacted pain reduction during the VR experience. The internal reliability of the PCS-C in the current sample was high (Cronbach’s α = 0.94). This questionnaire was completed by patients prior to the VR session.

#### 2.2.3 Anxiety Sensitivity

The Child Anxiety Sensitivity Index (CASI) is a validated, 18-item self-report survey designed to measure how patients perceive symptoms of anxiety, with total scores ranging from 18-54^52^. Like the PCS-C, the CASI was used to determine if trait levels of anxiety sensitivity impacted pain reduction during the VR experience. This survey has also been used in other studies with VR in adolescents^37^. The internal reliability of the CASI in the current sample was good (Cronbach’s α = 0.87). This questionnaire was completed by patients prior to the VR session.

#### 2.2.4 Patient Experience

Patient experience was measured with a 14-item questionnaire where participants ranked how much they agreed with statements on a 4-point Likert scale. A score of 1 was equivalent to the statement of “Strongly agree;” scores of 2, 3, and 4 corresponded to “Agree,” “Disagree,” and “Strongly disagree,” respectively. Example statements included: *“Virtual Reality made it easier for me to tolerate my pain;” “I would recommend friends or family to try virtual reality;” “I felt calmer and less anxious after having used virtual reality;”* and “I *felt like I was visiting the places in the displayed environment.”* The parent(s) of participants were also asked to fill-out a similar survey to better understand their perspective of their child’s experience with VR.

### 2.3 VR Device and Distraction Methods

During VR-D sessions, all patients used the Starlight Xperience VR device (https://www.starlight.org/virtual-reality/), a commercially available headset supplied by the Starlight Xperience program from the Starlight Children’s Foundation. The Starlight VR device is a customized version of the standalone Lenovo Mirage Solo with Daydream VR headset. A set of integrated headphones delivers audio to the user to create a fully immersive experience. By moving their head and using a small handheld controller, patients interact and navigate within the VR environment. During the session, patients selected from multiple VR experiences and games including: “INVASION!,” “ASTEROIDS!,” “Ocean Rift,” “Hello Mars,” “Pebbles the Penguin,” “Space Pups,” “Asteroid Miner,” “Wonderglade,” “Baskhead,” “LEGO BrickHeadz,” “Bait!,” “Along Together,” “Flutter VR,” “Lola and the Giant,” “Mekorama VR,” and “Expeditions.” These experiences and games are simple, non-violent, and easy to play, making them appropriate and engaging for the pediatric population; they all provide a similar distraction-based VR experience.

### 2.4 Procedures

After determining eligibility and obtaining consent (and assent where appropriate), patients completed questionnaires for trait catastrophizing (PCS-C) and anxiety sensitivity (CASI). Following completion of the surveys, patients were asked to rate their current pain intensity, pain unpleasantness, and anxiety levels with the NRS scale before VR to obtain baseline values.

Following baseline assessments, patients were oriented to the VR headset and given a tutorial on how to use the device. They were also instructed to remove the headset should discomfort or nausea ensue. They were then allowed to engage in any one of the multiple preloaded experiences listed above and underwent a single session. The specific experience chosen and the length of time within that experience were recorded for analysis. After completing a maximum of 20 minutes of VR-D (mean VR experience time = 14.6 ± 4.46 minutes), the VR device was removed. Pain intensity, pain unpleasantness, and anxiety were recorded immediately after completing the VR session and again at 15 minutes and 30 minutes after the experience. Patients completed a 14-item experience questionnaire after completing the VR session. Parents were given a similar survey. Figure 1 demonstrates the study design.

### 2.5 Statistical Analysis

All statistical analyses were done in SAS 9.4 (the SAS Institute, Cary, NC). A *p* value of 0.05 was used as the cutoff for statistical significance. Statistical significance with Bonferroni adjustment for multiple comparison for the primary outcome (change from baseline in pain intensity at 3 individual time points after VR-D) was assessed. AR(1) – first-order autoregressive covariance structure was used in all mixed effects models.

#### 2.5.1 Descriptive Analysis

Descriptive statistics were calculated and summarized for all baseline variables and change from baseline for outcome variables. Mean and standard deviation and/or median and interquartile range (IQR) were used for continuous variables. Frequency and percentage were used for categorical variables. For graphical purposes, mean pain and anxiety ratings are presented in Figure 2.

**Figure 2.**
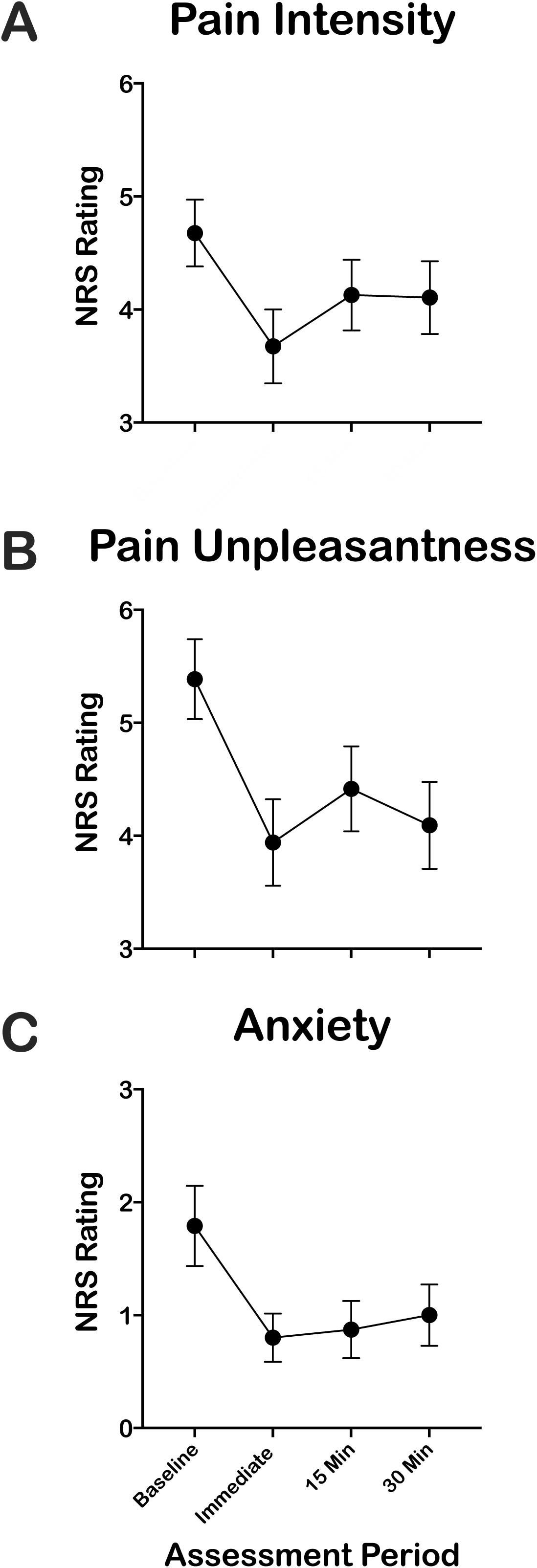
Ratings of Pain and Anxiety before and following VR-D. Pain intensity (A), pain unpleasantness (B), and anxiety (C) were collected before and up to 30 minutes following VR-D using the NRS. All patients (n = 50) provided an NRS score at baseline and immediately after VR-D. NRS scores were collected in 47 (94%) and 38 (76%) patients at 15- and 30-minutes following VR-D, respectively. This figure is provided for graphical purposes only.

#### 2.5.2 Changes in Pain and Anxiety following VR-D

Pain intensity, unpleasantness, and anxiety after VR-D were compared to those at baseline using paired tests (t-test or signed-rank, as appropriate) at individual time points. Changes from baseline were analyzed with mixed effects models where time (immediately, 15-, or 30-minutes after VR-D) was used as a categorical fixed effect, with and without adjustment for other covariates (patient age, sex, race, ASA, and postoperative day - POD), as well as baseline pain catastrophizing and anxiety.

#### 2.5.3 Associations in Baseline Outcomes

To test the association between baseline pain catastrophizing and anxiety sensitivity with baseline outcomes (pain intensity, pain unpleasantness, and anxiety), Pearson or Spearman correlation coefficients were derived as appropriate.

#### 2.5.4 Impact of Psychological Factors on Changes in Pain and Anxiety

Mixed effects models were used to examine the above associations and the effect of PCS-C and CASI on changes from baseline of the outcome NRS variables, where time (immediately, 15-, or 30-minutes after VR-D) was used as a categorical fixed effect, with and without adjustment for other covariates (patient age, sex, race, ASA status, and POD).

#### 2.5.5 Missing Data

Missing data was examined, and all available data were used in the statistical analysis.

## 3. Results

### 3.1 Baseline Participant Characteristics

We enrolled a total of 50 patients over 7 months. All patients (n = 50) completed the pain and anxiety assessment at baseline and immediately following the session: 94% (n = 47) completed the 15-minute assessment and 76% (n = 38) completed the 30-minute assessment. The missing data resulted from limitations in the clinical environment, including patients leaving for imaging studies, patients being seen by or receiving care from the care team, or patients falling asleep. Table 1 displays the study population demographics and scores of baseline measures. Patients were mostly adolescents, male and Caucasian. Of the 50 patients recruited, 19 (38%) underwent abdominal surgery, 17 (34%) underwent Nuss repair of pectus excavatum or chest surgery, and 14 (28%) underwent orthopedic procedures (such as posterior spinal fusion or major hip surgery). Most of the patients had an ASA status of I or II, so status was combined to reflect low (ASA I and II) and moderate/severe (ASA III and IV) levels of disease status. Most of the patients recruited were in the low disease group. Patients reported moderate levels of pain intensity and unpleasantness but low levels of anxiety prior to VR-D. Patients had moderate levels of pain catastrophizing on the PCS-C and average levels of anxiety sensitivity on the CASI ^51^. Patients with higher pain catastrophizing scores had higher pain intensity (Spearman correlation coefficient = 0.49, *p* = 0.0004) and unpleasantness (Spearman correlation coefficient = 0.45, *p* = 0.001) NRS scores prior to the VR experience (Supplementary Table 1). Anxiety sensitivity (CASI) was not associated with pain intensity and unpleasantness scores. Neither pain catastrophizing nor anxiety sensitivity were associated with anxiety NRS scores.

**Table 1.** Demographic, Survey, and Medical Data from Study Participants. Abbreviations: PCS-C, Pain Catastrophizing Scale for Children; CASI, Child Anxiety Sensitivity Index; ASA, American Society of Anesthesiologists.

| **Variable** | **Mean (SD) or**<br>**Number (%)** |
| --- | --- |
| **Age (years)** | 15.6 (2.5) |
| **Sex** |  |
| *Males* | 31 (62%) |
| *Females* | 19 (38%) |
| **Race** |  |
| *Caucasian* | 44 (88%) |
| *Non-Caucasian* | 6 (12%) |
| **Surgery Type** |  |
| *Abdominal* | 19 (38%) |
| *Pectus/Chest* | 17 (34%) |
| *Orthopedic* | 14 (28%) |
| **ASA Physical Status** |  |
| *I/II (Healthy, Mild Systemic Disease)* | 29 (58%) |
| *III/IV (Severe or life-threatening Disease)* | 21 (42%) |
| **Baseline NRS Scores** |  |
| *Pain intensity (0-10)* | 4.68 (2.09) |
| *Pain unpleasantness (0-10)* | 5.39 (2.50) |
| *Anxiety (0-10)* | 1.79 (2.52) |
| **Psychological Factors** |  |
| *Catastrophizing (PCS-C)* | 17.6 (11.8) |
| *Anxiety (CASI)* | 30.3 (6.7) |

### 3.2 Primary Outcome - Pain Intensity

There were moderate to large effects of VR-D on pain intensity (Figure 2A). Wilcoxon signed-rank test (Table 2) showed that pain intensity decreased immediately (Median (IQR) = -1.00 (-2.00, 0), *p* < 0.0001) following the session. This change remained significant at 15 minutes (Median (IQR) = 0 (−1.00, 0), *p* = 0.02). Change in pain intensity following the VR-D session was no longer significant at 30 minutes. The mixed effects model with adjustment for covariates showed that reduction in pain intensity was statistically significant compared to baseline when assessed immediately (LSM (SE) = -1.3 (0.3), *p* < 0.0001) after VR-D (Table 3), but not at 15 or 30 minutes. The decrease immediately after VR-D remained significant after adjustment for multiple comparisons.

**Table 2.** Mean (SD) and Median (IQR) Changes in Pain Intensity, Pain Unpleasantness, and Anxiety NRS Scores Following VR-D. Notes: Wilcoxon signed-rank test was used to compared changes. Abbreviations: ^NS^, Not significant; * *p*<0.05; ** *p* <0.01; *** *p*<0.001; **** *p*<0.0001

| **NRS Outcome following VR-D** | **N** | **Mean (SD)** | **Median (IQR)** |
| --- | --- | --- | --- |
| **Pain Intensity** |  |  |  |
| *Immediate* **** | 50 | -1.00 (1.14) | -1.00 (-2.00, 0) |
| *15 Minutes* * | 47 | -0.40 (1.12) | 0 (-1.00, 0) |
| *30 Minutes* <sup>NS</sup> | 38 | -0.33 (1.38) | 0 (-1.00, 0) |
| **Pain Unpleasantness** |  |  |  |
| *Immediate* **** | 50 | -1.45 (1.63) | -1.00 (-2.50, 0) |
| *15 Minutes* *** | 47 | -0.79 (1.53) | -1.00 (-2.00, 0) |
| *30 Minutes* *** | 38 | -1.09 (1.97) | -1.00 (-2.00, 0) |
| **Anxiety** |  |  |  |
| *Immediate* **** | 50 | -0.99 (2.00) | 0 (-1.00, 0) |
| *15 Minutes* ** | 47 | -0.97 (2.14) | 0 (-1.50, 0) |
| *30 Minutes* <sup>NS</sup> | 38 | -0.43 (1.56) | 0 (-1.00, 0) |

### 3.3 Secondary Outcome - Pain Unpleasantness

VR-D had a moderate to large effect on pain unpleasantness (Figure 2B). Wilcoxon signed-rank test (Table 2) revealed that pain unpleasantness decreased immediately following the VR-D session (Median (IQR) = -1.00 (−2.50, 0), *p* < 0.0001); this change remained significant at both 15 (Median (IQR) = -1.00 (−2.00, 0), *p* = 0.0008) and 30 (Median (IQR) = -1.00 (−2.00, 0), *p* = 0.0001) minutes following session completion. In our adjusted mixed effects models, reductions in pain unpleasantness were observed immediately (LSM (SE) = -1.42 (0.2), *p* < 0.0001), 15 minutes (LSM (SE) = -0.8 (0.2), *p* = 0.0035), and 30 minutes (LSM (SE) = -1.10 (0.3), *p* = 0.0005) following VR-D (Table 3).

**Table 3.** Mixed-Effect Model Reflecting Changes in Pain Intensity, Pain Unpleasantness, and Anxiety NRS Scores Following VR-D. Mixed Effects Models were run without (unadjusted) and with (adjusted) covariates. Notes: ^α^, Study covariates including demographic (age, sex, race), the time following surgery (postoperative day, POD), ASA status, and overall scores on the PCS and CASI. Abbreviations: ^NS^, Not significant; * *p*<0.05; ** *p* <0.01; *** *p*<0.001; **** *p*<0.0001

| **NRS Outcome following VR-D** | **Unadjusted Model**<br>**LSM (SE)** | **Adjusted Model<sup>α</sup>**<br>**LSM (SE)** |
| --- | --- | --- |
| **Pain Intensity** |  |  |
| *Immediate* | -1.0 (0.2) **** | -1.3 (0.3) **** |
| *15 Minutes* | -0.4 (0.2) * | -0.5 (0.3)<sup>NS</sup> |
| *30 Minutes* | -0.3 (0.2)<sup>NS</sup> | -0.5 (0.3)<sup>NS</sup> |
| **Pain Unpleasantness** |  |  |
| *Immediate* | -1.4 (0.2) **** | -1.8 (0.4) **** |
| *15 Minutes* | -0.8 (0.2) ** | -1.1 (0.4) ** |
| *30 Minutes* | -1.1 (0.3) **** | -1.4 (0.4) *** |
| **Anxiety** |  |  |
| *Immediate* | -1.0 (0.3) **** | -0.9 (0.5)<sup>NS</sup> |
| *15 Minutes* | -1.0 (0.3) ** | -0.9 (0.5)<sup>NS</sup> |
| *30 Minutes* | -0.6 (0.3) * | -0.6 (0.5)<sup>NS</sup> |

### 3.4 Secondary Outcome - Anxiety

We found moderate to large effects of VR-D on anxiety (Figure 2C). Wilcoxon signed-rank test (Table 2) revealed anxiety decreased from baseline immediately following the session (Median (IQR) = 0 (−1.00, 0), *p* < 0.0001). Anxiety scores remained lower at 15 minutes following the VR-D session (Median (IQR) = 0 (−1.50, 0), *p* = 0.0014). This change was no longer significant at 30 minutes following the VR-D session. However, based on the mixed effects model with adjustment for covariates, no significant changes were observed for anxiety immediately and following 15 and 30 minutes (Table 3).

### 3.5 Effects of Psychological Factors on Changes in Scores

Using a series of adjusted-mixed effects models, levels of trait pain catastrophizing or anxiety sensitivity were not associated with changes in pain and anxiety NRS scores following VR-D (Table 4).

**Table 4:**
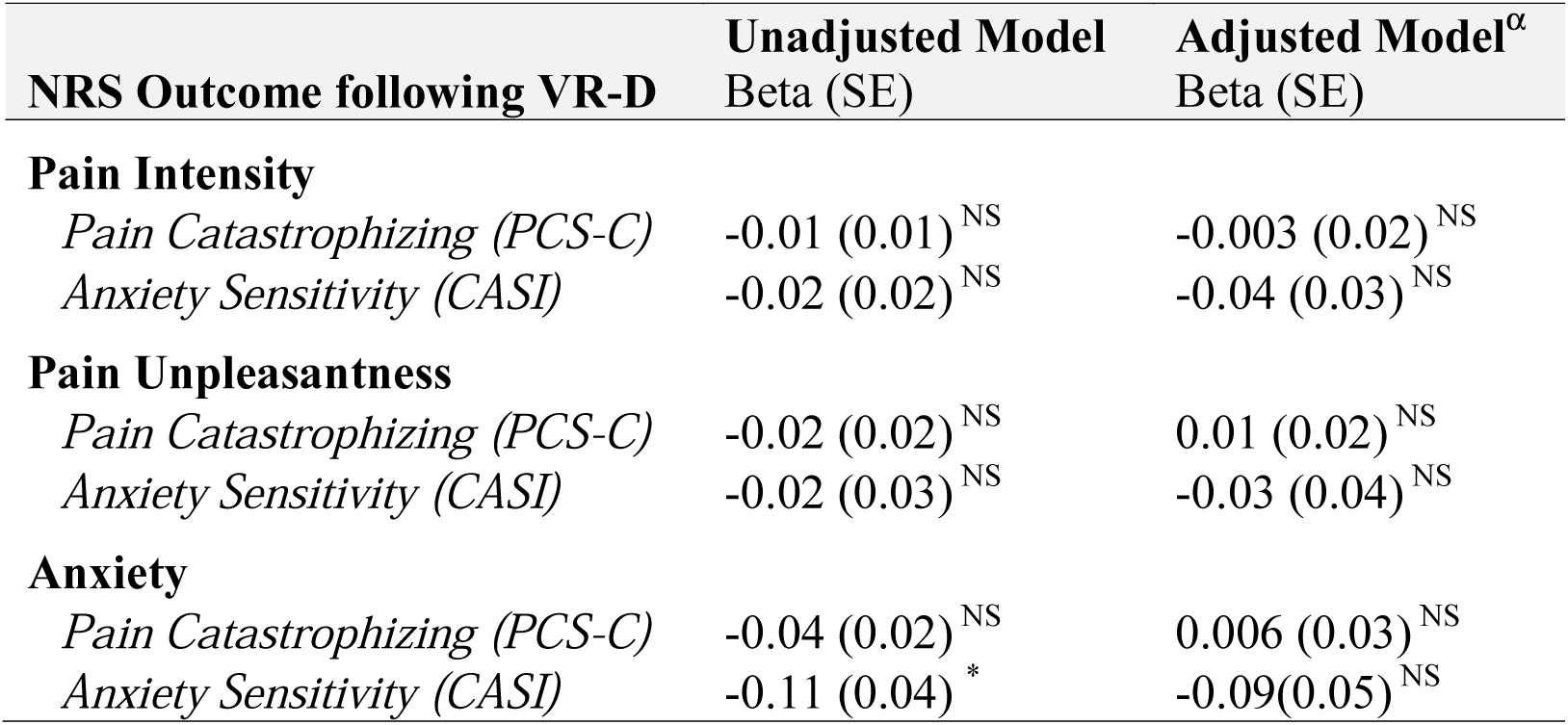
Impact of Psychological Factors on Changes in Pain Intensity, Pain Unpleasantness, and Anxiety NRS Scores Following VR-D. Mixed Effects Models were run for PCS-C and CASI without (unadjusted) and with (adjusted) other covariates. Notes: ^α^, Study covariates including demographic (age, sex, race), the time following surgery (postoperative day, POD), ASA status, and controlling for the overall scores on the PCS or CASI. Abbreviations: ^NS^, Not significant; * *p*<0.05

| **NRS Outcome following VR-D** | **Unadjusted Model**<br>**Beta (SE)** | **Adjusted Model<sup>α</sup>**<br>**Beta (SE)** |
| --- | --- | --- |
| **Pain Intensity** |  |  |
| *Pain Catastrophizing (PCS-C)* | -0.01 (0.01)<sup>NS</sup> | -0.003 (0.02)<sup>NS</sup> |
| *Anxiety Sensitivity (CASI)* | -0.02 (0.02)<sup>NS</sup> | -0.04 (0.03)<sup>NS</sup> |
| **Pain Unpleasantness** |  |  |
| *Pain Catastrophizing (PCS-C)* | -0.02 (0.02)<sup>NS</sup> | 0.01 (0.02)<sup>NS</sup> |
| *Anxiety Sensitivity (CASI)* | -0.02 (0.03)<sup>NS</sup> | -0.03 (0.04)<sup>NS</sup> |
| **Anxiety** |  |  |
| *Pain Catastrophizing (PCS-C)* | -0.04 (0.02)<sup>NS</sup> | 0.006 (0.03)<sup>NS</sup> |
| *Anxiety Sensitivity (CASI)* | -0.11 (0.04)<sup>*</sup> | -0.09(0.05)<sup>NS</sup> |

### 3.6 Study Covariates

When assessing the impact of other covariates on the effect of VR-D, mixed effects models showed that older participants had less decrease in anxiety (Beta (SE) = 0.28 (0.11), *p* = 0.0117), pain unpleasantness (Beta (SE) = 0.17 (0.08), *p* = 0.0378), and pain intensity (Beta (SE) = 0.12 (0.06), *p* = 0.0439). Female participants had a greater decrease in pain unpleasantness compared to male participants [Difference in LSM (95% CI) = 0.80 (0.01, 1.58), *p* = 0.0465]. No other variables (ASA, POD, or race) impacted the effect of VR-D.

### 3.7 Patient Satisfaction and Experience with Virtual Reality

Patients reported a very positive overall experience with VR. When asked if they would recommend VR to friends and family, 94% of children either strongly agreed (64.6%) or agreed (29.2%). A total of 42 patients (87.5%) believed that they felt “calmer and less anxious after having used VR,” and the same number of patients (87.5%) believed that VR “made it easier for [them] to tolerate [their] pain.” Parents also reported a positive overall experience when asked the same questions. A total of 38 parents (95%) who completed the experience questionnaire felt that they strongly agreed (83%) or agreed (13%) that they would recommend VR to friends or family. A total of 36 (90%) believed that VR made their child feel calmer and less anxious and 36 (90%) believed that VR helped their child tolerate pain.

## 4. Discussion

The results of this preliminary study demonstrate that a single session of VR-D transiently decreased pain intensity, pain unpleasantness, and anxiety in children and adolescents with acute postoperative pain after major surgery. However, these reductions in pain and anxiety were not sustained. Children with higher pain catastrophizing scores had higher levels of both pain intensity and pain unpleasantness at baseline, but they did not experience a larger reduction in pain in response to VR-D as compared to children with lower levels of pain catastrophizing. Anxiety sensitivity did not influence VR-D’s ability to reduce pain intensity, pain unpleasantness, nor anxiety.

Current postoperative pain management strategies center around pharmacological management, primarily with opioids^8^. While opioid exposure can increase the risk of persistent use^12^, inadequate pain management is also problematic. Ineffective management of postoperative pain results in poor surgical outcomes, delayed wound healing, and increased likelihood of developing chronic pain^5, 6^. Despite widespread use of multimodal analgesia, the percentage of patients experiencing severe pain after surgery has not changed over the last 20 years^1, 2^. It is therefore imperative to consider alternative, innovative strategies to provide safe, effective, nonpharmacologic therapies to children and adolescents after surgery to optimize pain management while minimizing opioid use^13^. VR can be considered a safe, effective tool with this potential.

Our results are consistent with previous literature demonstrating the effectiveness of VR in treating pain. The use of VR-D to help minimize pain has been a topic of interest for several years, beginning with its use to help manage pain in adult and pediatric patients with burns^15-18,23, 26, 27, 32, 34-36, 39, 41, 53-55^. Dating back to the early 2000s, one of the first groups studying this topic used SnowWorld, a VR program designed to help control pain during dressing changes in burn patients^34^. The authors noted a 41% decrease in pain while the patients were exposed to the VR condition as compared to control patients; they also noted a strong negative correlation between VR immersion and pain ratings^34^. A similar study assessing the role of VR in burn patients saw a 44% reduction in pain-related cognition (time spent thinking about pain), a 32% reduction in affective pain (emotional unpleasantness associated with pain), and a 27% reduction in sensory pain along with positive feelings about a VR intervention^17^. Similarly, Carrougher and colleagues found similar results in pediatric burn patients: a 31% reduction in pain unpleasantness, a 41% reduction in cognitive pain, and a 27% reduction in the worst pain experienced while patients were immersed in a VR environment^15^.

Researchers have also investigated the role of VR-D in minimizing pain during minor procedures, such as while obtaining IV access^21^; during dental procedures^19,22^; during labor^56, 57^; and even during urologic procedures^58^. However, only a few studies address the use of VR to help manage acute postoperative pain^59, 60^, and no study to date has addressed this topic in children. While it is clear that children will often require opioid analgesics to assist with postoperative pain control, VR can be useful as a complementary adjunct to opioid medications^28^. In 2007, Hoffman and colleagues evaluated the effect of using VR-D versus VR-D plus opioids in treating pain using functional magnetic resonance imaging (fMRI)^28^. Brain activity patterns supported the analgesic benefit of using VR-D in combination with opioid therapy, supporting the incorporation of VR-D into multimodal analgesia^28^.

While our results confirm the results of previous studies, our results are also unique, as they expand the potential role of VR-D in pain management therapy. The purpose of our study was to preliminarily assess the ability of VR-D to transiently decrease acute postoperative pain after a single session and to determine whether anxiety sensitivity may influence the efficacy of VR-D to help manage pain in a broad, pediatric population experiencing significant pain after surgery. This small, single VR session study sets the stage for the development of a larger, randomized clinical trial assessing the impact of VR-D on acute postoperative pain and anxiety versus control, as well as a study to explore the use of an increased number of VR-D sessions in the immediate postoperative period. Furthermore, we will also explore the impact that VR-D may have on postoperative opioid consumption. These larger trials will help us understand how VR can help decrease pain and minimize use of pharmacological agents thereby minimizing the risk of both persistent pain and persistent opioid use. Our patient experience data demonstrates that most patients enjoyed the VR experience and would recommend it to friends. Patients felt that the VR session improved their ability to tolerate pain and helped with their anxiety. Positive patient experiences suggest that patients would likely be compliant with VR treatment, further supporting its utility and viability in the postoperative arena.

There were several limitations to this study. First, the setup of this study did not allow for the comparison of VR-D against another distraction-based method to treat postoperative pain or a control of no additional treatment. We also were unable to assess the impact of VR-D on opioid consumption. The goal of this study was to test feasibility and obtain pilot data demonstrating the ability to use VR-D in acute postoperative patients while in the hospital and to demonstrate transient reductions in pain and anxiety. We recognize that a single VR-D session is unlikely to demonstrate a sustained effect on pain and anxiety in this population and plan to pursue further clinical studies in which patients engage in repeated sessions. Second, VR is a relatively new technology being integrated into the healthcare setting that requires instruction and experience for patients to become familiar with the device. Few patients had experience using a VR headset prior to participating in this study. As a result, some patients had difficulty navigating menus and games which may have negatively impacted their overall VR experience. In future studies, we will incorporate targeted education using a standardized script to allow patients to feel familiar and comfortable with the device prior to the study and will dedicate a session to device and program orientation. Third, our VR session was only a single session incorporated during the postoperative period. It is quite possible that increased length of distraction duration or repeated experiences over several days could have augmented the results of this experience as the patients became more familiar with the device and subsequently more immersed, resulting in increased and sustained benefit. Future study will certainly incorporate these aspects. Fourth, this study used a diverse patient population. While this is a limitation in the sense that our population was not standardized, each patient served as his or her own control (baseline), eliminating the issue of patient diversity in the sample. Furthermore, by using a diverse patient population, we were able to show that VR can be used successfully in many different types of patients thereby expanding the generalizability of these findings and future study design. Finally, there is also the potential that patients had bias when self-reporting pain scores. If they expected the treatment to make an impact, they may have self-reported lower pain and anxiety scores following their VR experience. Such bias will be addressed in future study with addition of a control arm.

In summary, our study demonstrates the successful extension of VR-D into the pediatric, acute postoperative setting for pain management. VR-D provides a transient and minor decrease in pain and anxiety after a single session. Future research is needed to investigate alternative applications of VR in the postoperative setting with the goal of providing greater and more sustained pain and anxiety relief. One potential novel approach is to incorporate established mind-body therapies with VR, allowing for greater, more sustained reductions in pain and anxiety. Without VR, distraction alone provides little lasting benefit in pain management or significant reductions in pain. However, while VR-D is more effective than traditional modes of distraction, VR for pain management will likely require extension of pain relief strategies beyond distraction alone.

## Data Availability

Data will be available upon request.

## Acknowledgements

The authors thank Maria Ashton MS, RPH, MBA for providing writing assistance, editing and proofreading.

---

**Supplementary Table 1:** Spearman Correlation Between Baseline NRS Scores and Surveys. Abbreviations: ^NS^, Not significant; ** *p*<0.01, *** *p*<0.001

| Baseline NRS | Pain<br>Catastrophizing<br>(PCS-C) | Anxiety<br>Sensitivity<br>(CASI) |
| --- | --- | --- |
| Pain Intensity | 0.49<sup>***</sup> | 0.17<sup>NS</sup> |
| Pain Unpleasantness | 0.45<sup>**</sup> | 0.20<sup>NS</sup> |
| Anxiety | -0.04<sup>NS</sup> | 0.17<sup>NS</sup> |

